# Estimating the effectiveness of syndromic screening at airports for Bundibugyo ebolavirus disease

**DOI:** 10.64898/2026.06.11.26355442

**Authors:** Billy J Quilty, CMMID Bundibugyo Working Group

**Affiliations:** Charité – Universitätsmedizin Centre for Mathematical Modelling of Infectious Diseases, London School of Hygiene & Tropical Medicine

## Abstract

We used a stochastic simulation model to estimate the effectiveness of exit and/or entry syndromic screening at airports for Bundibugyo ebolavirus disease (BVD). For a 12-hour international flight from DRC or Uganda at 86% screening sensitivity, approximately 73% of infected people presenting for travel would evade combined exit and entry screening (95% CrI: 68–77%). Among those who successfully boarded, approximately 92% would arrive without detection (95% CrI: 90–94%). This high undetected fraction reflects the relative durations of the incubation period (approximately 7.7 days; Wamala et al. 2010^1^) and the onset-to-severe-disease interval (approximately 4 days; Funk & Abbott 2026^2^): most infected travellers board before symptom onset and are undetectable by any syndromic screen, whilst those who are symptomatic progress rapidly to illness severe enough to preclude travel. Because the proportion of confirmed cases presenting without fever is itself uncertain, we also report a conservative scenario in which all confirmed cases are febrile (0% asymptomatic-equivalent), under which the undetected fraction remains substantial at approximately 64% (95% CrI: 57–70%). This is compounded during active epidemic growth, when recently exposed (and therefore pre-symptomatic) cases are overrepresented among travellers. Syndromic airport screening offers limited protection against BVD spread via air travel, and should be complemented by outbreak control at source, strengthened clinical surveillance in receiving countries with high travel connectivity to affected areas, and post-departure self-monitoring guidance for travellers from affected regions.

## 1 Introduction

Bundibugyo ebolavirus (BDBV) is one of six known ebolaviruses, previously identified in outbreaks in Uganda (2007)^3^ and the Democratic Republic of the Congo (DRC; 2012)^4^. An outbreak of Bundibugyo ebolavirus disease (BVD) affecting Ituri Province in DRC and with confirmed spread to Kampala, Uganda was declared a Public Health Emergency of International Concern (PHEIC) by WHO in May 2026^5^. As of 15 July 2026, the outbreak had spread across 46 health zones in five DRC provinces (Ituri, North Kivu, South Kivu, Haut-Uele, and Tshopo), with 2,124 confirmed cases and 828 deaths (case fatality ratio 39%) recorded in DRC, 20 confirmed cases and two deaths in Uganda, and single imported cases identified in France and Germany^6^; a basic reproduction number of 1.71 was estimated earlier in the outbreak from the confirmed case series^7^. Unlike previous Zaire ebolavirus outbreaks, no approved BVD-specific therapeutics or vaccines exist, and the outbreak is occurring against a backdrop of insecurity, high population mobility, and a large informal healthcare network. Together these features heighten concern about international spread via air travel.

Critically for screening effectiveness, clinical data from the 2026 outbreak indicate that fever is recorded at presentation in only approximately 74% of confirmed BVD cases, with haemorrhagic signs in just 10%; gastrointestinal symptoms (diarrhoea 67%, vomiting 67%) predominate instead^8^. This is towards the lower end of pooled estimates for Ebola virus disease more broadly (76%, 95% CI 662̆ 01385%;^9^). Since airport syndromic screening relies primarily on fever detection, this pattern of clinical presentation has direct implications for how many infected travellers such screening can realistically identify, as explored throughout this analysis.

Syndromic airport screening (thermal scanning, symptom questionnaires, and direct clinical assessment) was deployed extensively during the 2014–2016 West Africa Ebola epidemic and again during the 2018–2020 DRC outbreaks of Zaire ebolavirus^10^. Its effectiveness, however, is constrained by the natural history of infection: travellers who have been exposed but have not yet developed symptoms will not be febrile at the time of screening, and therefore will not be detected. Whether a traveller is symptomatic at departure or arrival depends on when they were exposed relative to their travel date, on the distribution of the incubation period (infection to symptom onset), and on the time from symptom onset to progression to severe, detectably ill disease.

For BVD specifically, three published incubation period estimates exist and were identified in a recent systematic review^11^. Wamala et al. (2010)^1^ reported a mean of 7.68 days (SD 3.53; 95% CI 7.04–8.32; n = 116) from the 2007 Uganda outbreak. MacNeil et al. (2010)^12^ reported 6.3 days (95% CI 5.2–7.3; n = 24 contacts with confirmed exposure dates) from the same outbreak. Kratz et al. (2015)^13^ reported 11.3 days (range 2–27; n = 3) from the 2012 Isiro DRC outbreak. We adopt the Wamala et al. (2010) estimate as our primary value, given its substantially larger sample size; the rationale for not pooling the three estimates, and how the resulting parameter uncertainty is propagated, is described in Methods. The MacNeil and Kratz estimates instead define the range explored in a dedicated incubation-period sensitivity analysis (Figure 4). Combined with a Bayesian reanalysis of the 2012 Isiro line list, which we conduct to estimate a mean onset-to-severe-disease interval of approximately 4 days (Funk & Abbott 2026^2^; see Methods), these two values anchor the natural-history assumptions underlying the model developed below.

Total flight durations from DRC or Uganda to international destinations typically range from approximately 10 to 16 hours depending on destination, measured as end-to-end travel time including layovers rather than direct airborne time. We model a 12-hour flight as a representative default whilst examining how screening effectiveness varies across a range of flight durations and epidemiological scenarios.

## 2 Methods

### 2.1 Simulation model

We used a stochastic individual-based simulation model to evaluate the probability that an infected traveller would be detected at (i) exit screening before departure, (ii) as severely ill during the flight, (iii) at entry screening on arrival, or (iv) remain undetected throughout.

For each simulated traveller, an incubation period *t*_inc_ and an onset-to-severe-disease interval *t*_inf_ were drawn independently from Gamma distributions. The time at which a traveller commenced their flight relative to their infection was drawn uniformly from [0, *t*_inc_ + *t*_inf_ + *t*_rec_], where *t*_rec_ is an individual-specific draw from the onset-to-death Gamma posterior (Funk & Abbott 2026^2^), used as a proxy for the time to resolution of infection (death or recovery) for all simulated travellers, defining the full window within which departure could occur. Travellers hospitalised before their scheduled departure were excluded.

Exit screening was assumed to detect any traveller who was symptomatic (*t*_flight departure_ > *t*_inc_) with probability *p*_exit_; travellers who became severely ill during the flight or remained symptomatic on arrival were subject to entry screening with probability *p*_entry_. A proportion of all infections was assumed to be undetectable by fever-based syndromic screening throughout — comprising both truly asymptomatic infections and symptomatic-but-afebrile presentations, since thermal/fever screening cannot detect either — and remained undetected throughout.

### 2.2 Parameters

Model parameters are listed in Table 1. The Isiro 2012 line list, the only publicly available BVD line list, lacks exposure dates and was re-analysed using doubly-censored Bayesian methods by Funk & Abbott (2026)^2^ to estimate Gamma-distributed onset-to-severe-disease and onset-to-death delays (onset-to-severe-disease posterior median shape 1.18, scale 3.41).

**Table 1:** Model parameters for the BVD default scenario.

| Parameter | Value | Source |
| --- | --- | --- |
| Flight duration | 12 h (default) | DRC/Uganda via connecting hub<br>(representative) |
| Exit screening sensitivity | 86% | Priest et al. 2011 (thermal image scanners) |
| Entry screening sensitivity | 86% | Priest et al. 2011 (thermal image scanners) |
| Proportion asymptomatic<br>(undetectable via fever<br>screening) | 25.7% | Derived as 1 fever prevalence at presentation<br>(Akilimali et al. 2026, 2026 DRC outbreak) |
| Incubation period mean | 7.68 days (95% CI: 7.048.32) | Wamala et al. 2010 (2007 Uganda BVD;<br>n=116) |
| Incubation period variance | 12.46 days<sup>2</sup> (SD = 3.53 days) | Wamala et al. 2010 |
| Incubation mean prior SD | 0.328 days (Wamala et al. SE<br>of the mean, SD/n) | Wamala et al. 2010 (SD 3.53, n=116); not<br>pooled with MacNeil/Kratz (see Introduction) |
| Onset severe disease mean | 4.03 days (95% CrI: 3.085.46) | Funk & Abbott 2026 posterior median |
| Onset severe disease<br>variance | 13.72 days<sup>2</sup> | Funk & Abbott 2026 posterior median |

As introduced above, we use the Wamala et al. (2010)^1^ estimate as the primary incubation-period value and propagate parameter uncertainty via a Normal prior on the mean (mean 7.68 days, SD 0.328 days — Wamala et al.’s own standard error of the mean, truncated above 0.5 days), scaling the corresponding variance to preserve the coefficient of variation (CV 0.46; Gamma shape 4.73 held constant across draws), consistent with treating relative incubation-period variability as a stable biological property. Because this prior reflects only within-study sampling uncertainty, the wider range spanned by the MacNeil and Kratz estimates (not pooled here, per the Introduction) is instead explored via the incubation-period sensitivity analysis (Figure 4).

Thermal and syndromic screening can detect a traveller only if they are febrile at the time of screening. Clinical data from the 2026 outbreak indicate that fever was recorded in only 74.3% of confirmed BVD cases at presentation^8^; the remaining 25.7% of symptomatic cases would never be detected by fever-based screening, regardless of scanner sensitivity or timing, and are therefore functionally equivalent to asymptomatic infection for the purposes of this model. We use this outbreak-specific, fever-negative fraction directly as the proportion asymptomatic parameter (25.7%), in place of a generic EVD-literature placeholder, since it is grounded in current-outbreak clinical data, though this estimate carries its own uncertainty (see Discussion, Limitations).

Real-world syndromic screening is not necessarily limited to thermal detection, however; visual observation of other signs and symptom questionnaires may also identify some afebrile but otherwise symptomatic travellers, so our estimates of the undetected fraction may be conservative in this respect.

### 2.3 Uncertainty propagation

For each of 1000 posterior draws sampled at random from the Funk & Abbott (2026)^2^ BVD posterior, we ran the simulation with 2,000 simulated travellers. On each draw, the onset-to-severe-disease delay was taken from the BDBV posterior (draw-specific Gamma mean and variance from the Funk & Abbott 2026 analysis), and the incubation period mean was drawn, independently of the onset-to-severe-disease delay rather than paired by draw index, from the Normal prior described above (Wamala et al.’s own standard error), with variance rescaled to preserve the coefficient of variation. Sampling these two sources of uncertainty independently means the reported credible intervals reflect their combined, uncorrelated variation rather than any assumed dependence between them.

### 2.4 Sensitivity analyses

Four sensitivity analyses were conducted, each addressing a source of uncertainty not captured by the primary posterior-propagation analysis above.

First, we examined how the proportion undetected varied across flight durations of 1 to 20 hours, retaining full posterior uncertainty on natural-history parameters.

Second, we explored the joint effect of the proportion of asymptomatic infection (0 to 60%) and the mean incubation period (1 to 14 days) on the proportion undetected for the default 12-hour flight, using posterior-median natural-history parameters; this addresses uncertainty in two parameters that are not themselves derived from the outbreak-specific posterior.

Third, to relax the baseline assumption of a stable epidemic, we re-ran the posterior analysis across epidemic doubling times of 4.5 to 44 days (spanning the 90% credible interval from the epiforecasts BVD 2026 growth-rate estimates^2^). In a growing epidemic, recently infected individuals are overrepresented among travellers relative to those infected further in the past, so the uniform exposure-time assumption underestimates the pre-symptomatic fraction. Following Gostic et al.^14,15^, we replaced the uniform draw on [0, *T*_max_] with a truncated-exponential distribution: each doubling time *T_d_* was converted to an exponential growth rate *r* = ln 2/*T_d_*, and the time since infection for each simulated traveller was drawn by inverse-CDF sampling, = ln(1 *U* (1 *e^r^ ^T^*^max^))/*r*, where *U* Uniform(0, 1) and *T*_max_ = *t*_inc_ + *t*_inf_ + *t*_rec_ as in the stable-epidemic case (see Simulation model). This weights the exposure distribution towards recent infections, as expected under exponential growth.

Fourth, we varied exit and entry screening sensitivity independently from 0 to 100% to quantify the marginal contribution of each screening layer.

### 2.5 Software

All analyses were performed in R (version 4.1; R Core Team 2024). The simulation model is implemented in the airportscreening R package, available at github.com/bquilty25/airport_screening_ebola_bvd. An interactive Shiny application for exploring screening parameters is included in the repository and can be launched locally by running airportscreening::run_app() after installing the package.

## 3 Results

### 3.1 Detection outcomes at 12 hours

For a 12-hour connecting flight from DRC or Uganda under the baseline scenario (86% screening sensitivity at both departure and arrival), we estimate that 21.1% (95% CrI: 16.9–25.5%) of infected travellers would be detected at exit screening, 0.6% (95% CrI: 0.2–1.2%) would become severely ill during the flight, and 5.4% (95% CrI: 4.1–6.7%) would be detected at entry screening, leaving 72.9% (95% CrI: 68.1–77.3%) of infected travellers undetected on arrival (Table 2). Under the theoretical best-case scenario (100% exit / 100% entry screening sensitivity), exit detection rises to 24.3% (95% CrI: 19.7–29.8%), yet 72.2% (95% CrI: 67.0–76.4%) of infected travellers would still arrive completely undetected due to boarding while pre-symptomatic or asymptomatic.

**Table 2:**
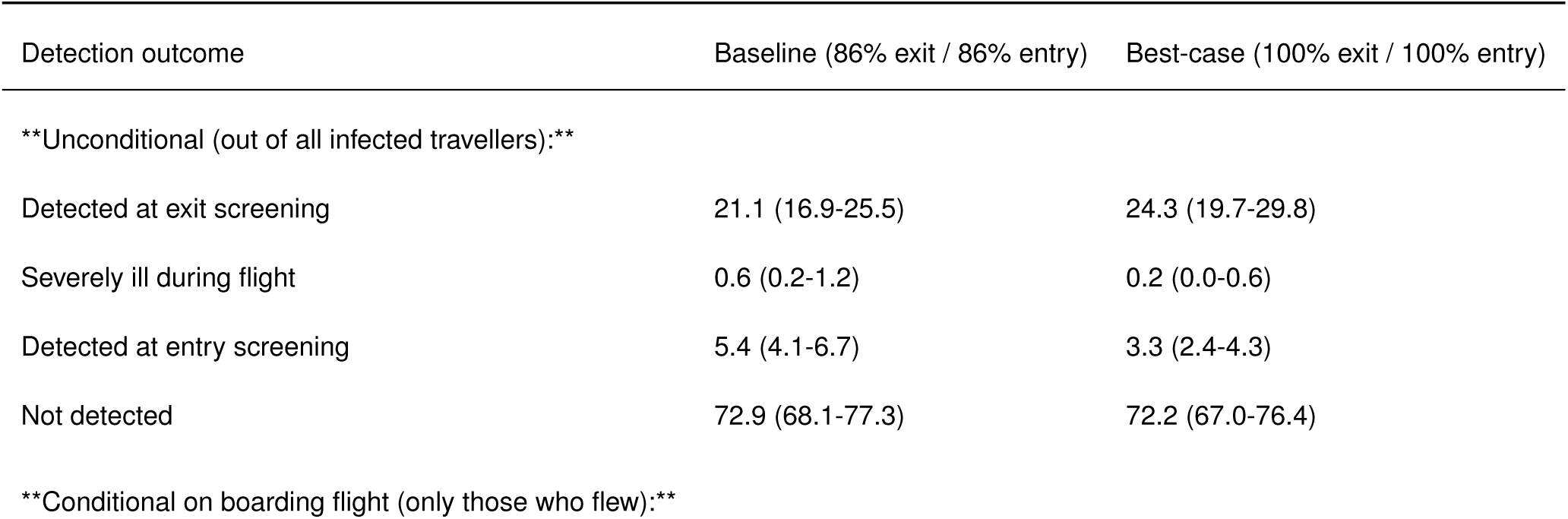
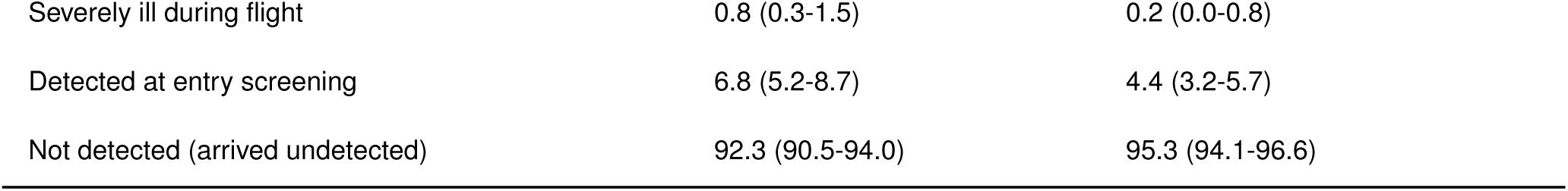
Estimated screening outcomes for 100 infected BDBV travellers on a 12-hour connecting flight from DRC or Uganda. Values are percentages (medians) with 95% credible intervals.

Of those who successfully boarded the flight (excluding those detected at exit screening), the proportion who arrived undetected was 92.3% (95% CrI: 90.5–94.0%) under baseline screening, and 95.3% (95% CrI: 94.1–96.6%) under the best-case (100% exit / 100% entry) scenario. Under the baseline scenario, 0.8% (95% CrI: 0.3–1.5%) of boarded cases became severely ill during the flight and 6.8% (95% CrI: 5.2–8.7%) were detected at entry screening; under the best-case scenario, 0.2% (95% CrI: 0.0–0.8%) became severely ill during flight and 4.4% (95% CrI: 3.2–5.7%) were detected at entry screening.

The natural-history distributions underlying these estimates are shown in Figure 1: the incubation-period distribution is narrow, reflecting the tight within-study prior described in Methods, whereas the onset-to-hospitalisation and onset-to-death distributions are wider, reflecting the limited information available in the small Isiro 2012 line list.

**Figure 1:**
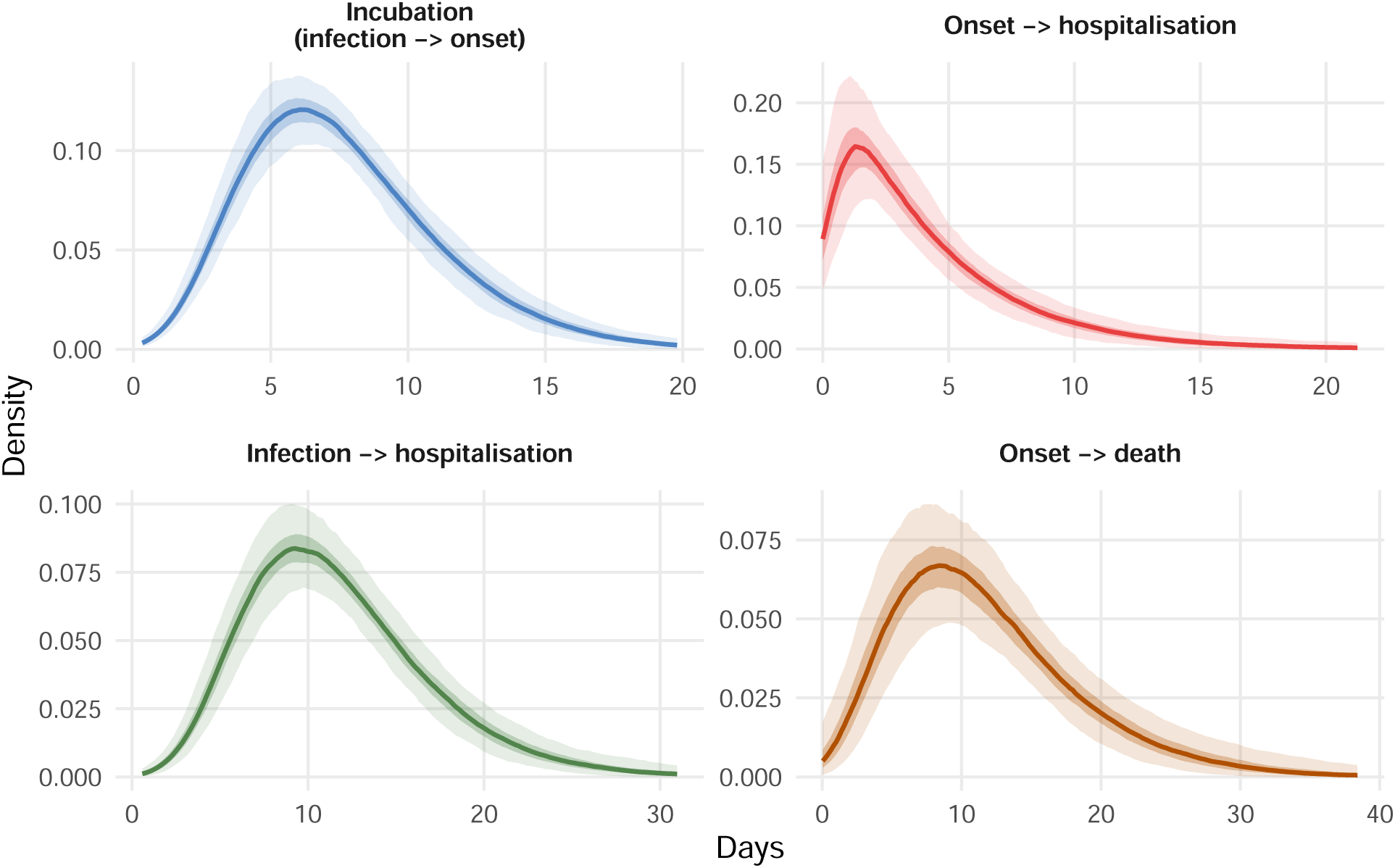
Posterior distributions of natural-history period durations for BVD. Each panel shows densities from 500 joint draws: the onset-to-hospitalisation and onset-to-death delays are drawn from the Bayesian BVD posterior (Funk & Abbott 2026^2^); the incubation period (infection to symptom onset) is drawn from a Normal prior on the mean (7.68 days, SD 0.328 days — Wamala et al.’s own standard error of the mean^1^) with CV preserved (Gamma shape fixed at 4.73). Shaded ribbons show 50% (dark) and 95% (light) pointwise credible bands across the 500 density curves; because the prior on the incubation mean reflects only within-study sampling uncertainty, the incubation-period ribbon is narrow relative to the other panels (see the incubation period sensitivity analysis below for structural uncertainty across the published range of estimates). The onset-to-hospitalisation distribution has shape ∼1.2 (nearly exponential), reflecting the limited information in the Isiro 2012 line list.

### 3.2 Detection outcome breakdown

Exit screening accounts for the largest share of detected cases; severely ill during flight and entry- detected cases together represent a small fraction, reflecting the short window between symptom onset and severe disease relative to the flight duration (Figure 2). This re-visualises the same 12-hour baseline scenario reported above in Table 2, decomposing it into its constituent detection pathways.

**Figure 2:**
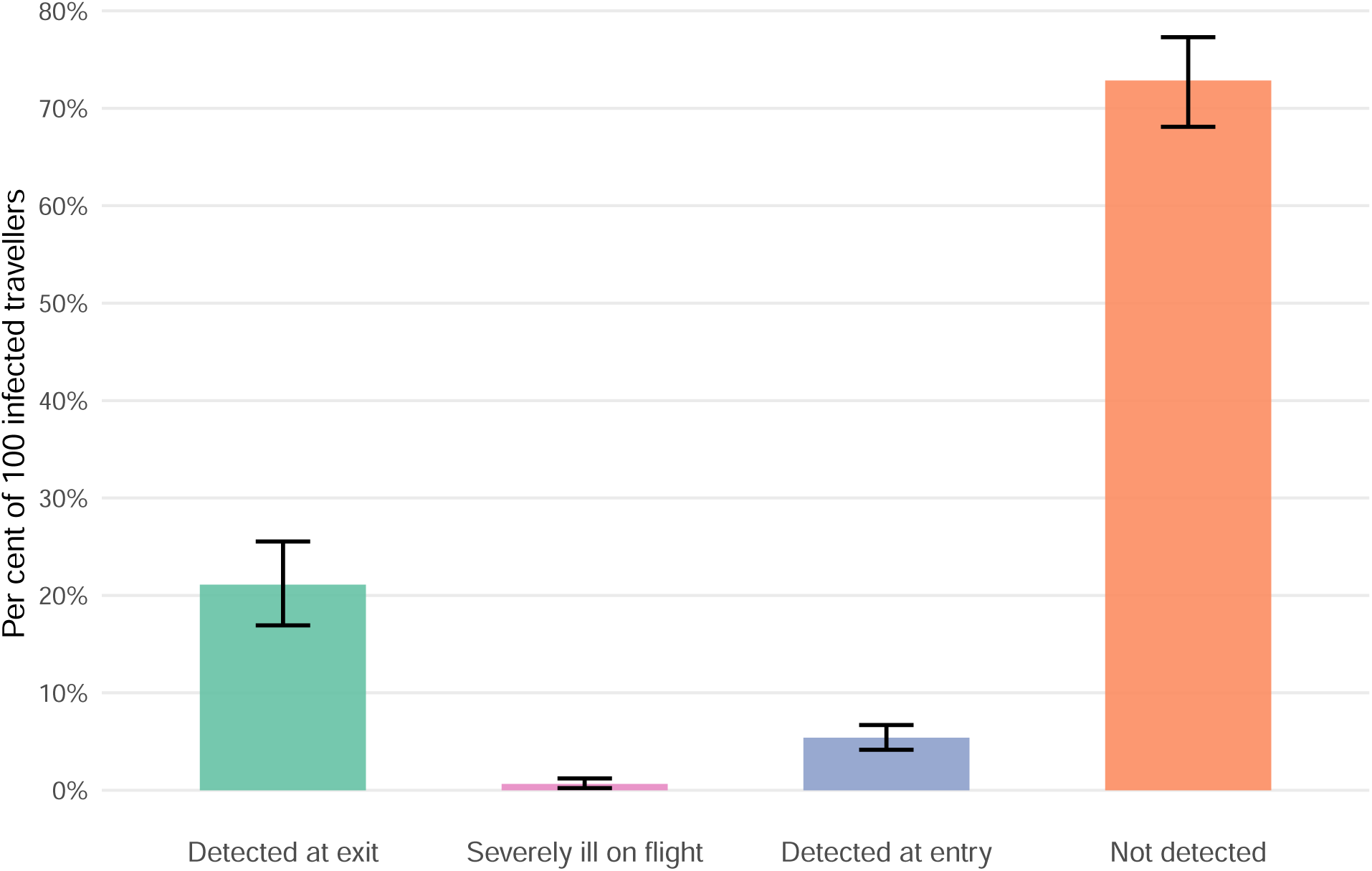
Estimated number of detected and undetected infected travellers per 100 infected travellers departing DRC or Uganda on a 12-hour connecting itinerary, stratified by detection outcome, with 95% credible intervals (whiskers).

### 3.3 Effect of flight duration on detection probability

As flight duration increases, the probability of detecting an infected traveller at entry screening rises owing to progression of disease during travel (Figure 3), though the overall undetected fraction remains high throughout. Even for a 20-hour flight (the longest typical single-airline connection), the proportion undetected remains 70.9% (95% CrI: 65.2–75.6%), little changed from the 12-hour default reported above.

**Figure 3:**
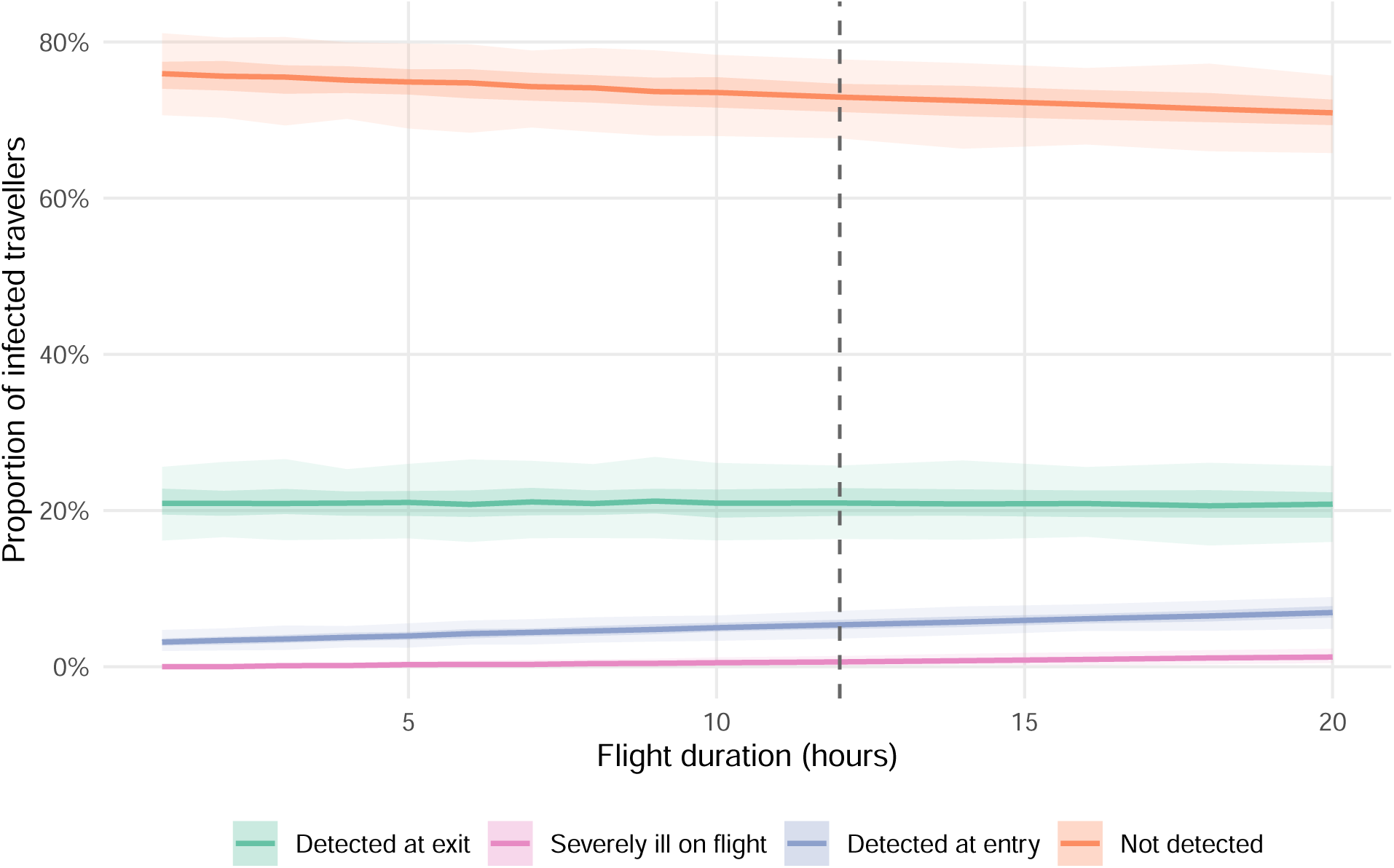
Detection probabilities as a function of flight duration, with uncertainty propagated across 1,000 posterior draws of the BVD onset-to-hospitalisation delay. Ribbons show 50% (dark) and 95% (light) credible intervals. The vertical dashed line marks 12 hours, the approximate DRC/Uganda connecting travel time to international destinations.

### 3.4 Sensitivity analysis: asymptomatic proportion and incubation period

The primary driver of the undetected fraction is the incubation period (Figure 4): even at our default afebrile/asymptomatic fraction of 25.7% (derived from 2026 outbreak fever-prevalence data; see Methods), the proportion undetected ranges from 36–82% across incubation periods of 1–14 days, with longer incubation periods allowing more travellers to depart before symptom onset. This fraction combines true asymptomatic infection, which is rare in EVD, with afebrile-but-symptomatic presentations that fail to trigger fever-based syndromic screening; the latter appear to be common for BVD based on current-outbreak clinical data^8^. If the afebrile/asymptomatic fraction were as high as 40%, the undetected fraction would rise to 49–86% across the same incubation range.

### 3.5 Sensitivity analysis: epidemic growth phase

The proportion of infected travellers going undetected increases as the epidemic doubling time shortens (Figure 5). If the true doubling time falls within the initial epiforecasts 90% credible interval (13.8–22.8 days), the model estimates a corresponding undetected fraction of approximately 76–77%. Under the more pessimistic latest-estimate lower bound (4.5 days), the undetected fraction rises to 85% (95% CrI: 82–88%), compared with 73% under a stable epidemic. This pattern is consistent with the general principle that screening performs less well during periods of rapid epidemic growth, when pre-symptomatic cases are overrepresented among travellers.

**Figure 4:**
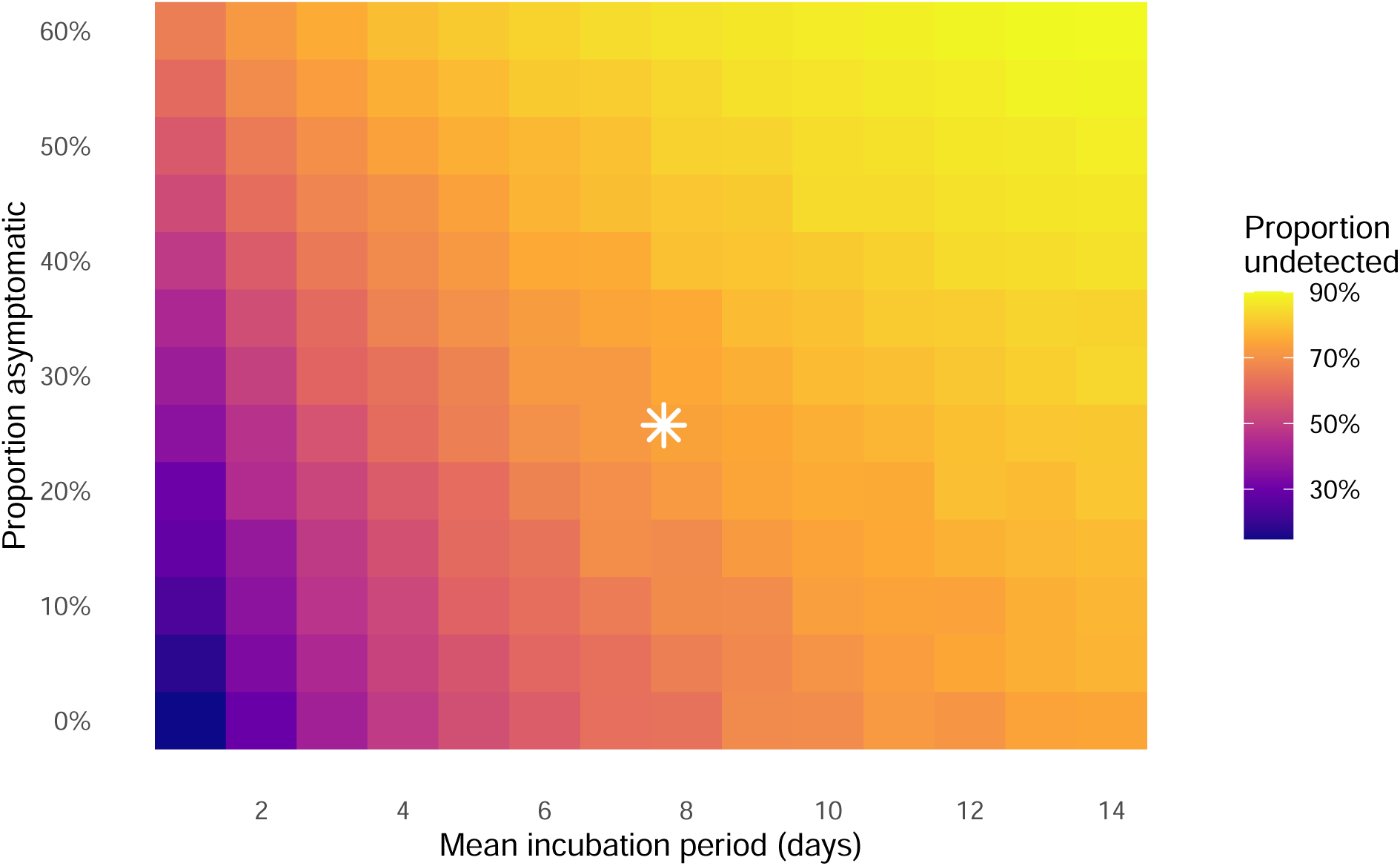
Sensitivity analysis: proportion of infected travellers undetected as a function of the mean incubation period (x-axis) and proportion of asymptomatic infection (y-axis), for a 12-hour flight with posterior-median BVD onset-to-hospitalisation parameters. As the mean incubation period varies, the variance is rescaled to preserve the coefficient of variation (CV = SD/mean 0.46, equivalent to a fixed Gamma shape of 4.73), consistent with the uncertainty propagation used in the main analysis. The star marks the primary BVD scenario (mean incubation 7.68 days, 25.7% asymptomatic). Full uncertainty propagation across the incubation period is shown in the main analysis.

**Figure 5:**
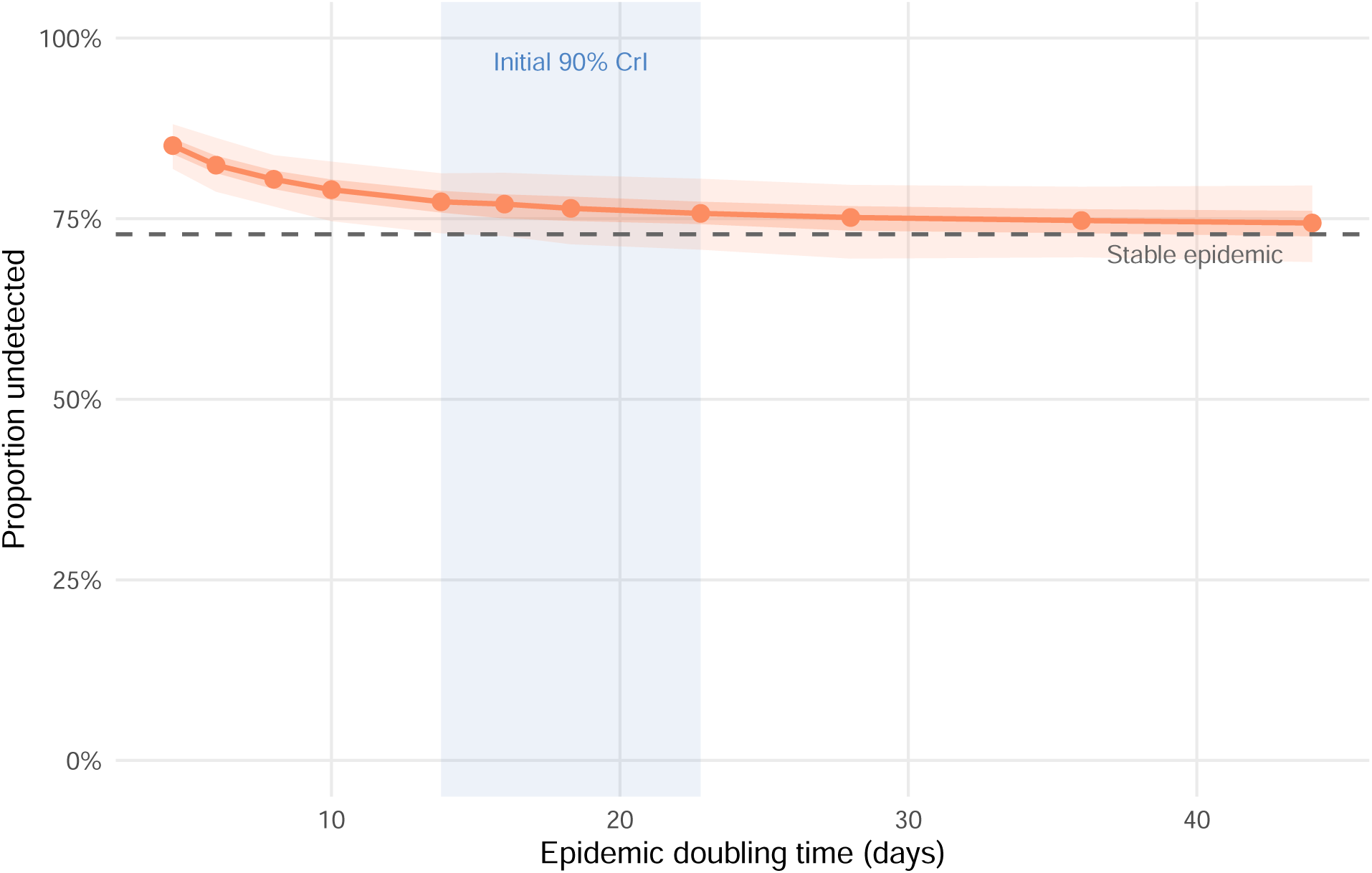
Effect of epidemic doubling time on the proportion of infected travellers going undetected. Each point shows the median across 500 posterior draws; ribbons show 50% (dark) and 95% (light) credible intervals. The shaded band marks the 90% credible interval for the initial doubling time of the current BVD outbreak (13.8–22.8 days; epiforecasts, 2026). The rightmost marker (dashed vertical) represents a stable epidemic (growth rate = 0; uniform exposure distribution).

### 3.6 Sensitivity analysis: screening sensitivity at exit and entry

Exit screening sensitivity has a larger marginal effect on the undetected fraction than entry screening sensitivity, because it acts on a larger at-risk window: any traveller already symptomatic at departure can be caught at exit, whereas entry screening only adds value for those who become symptomatic during the flight or who were missed due to imperfect screening at exit (Figure 6). When exit screening is absent entirely (sensitivity 0%) but entry screening is maintained at 86%, 74% of travellers remain undetected, compared with 79% when entry screening is absent and exit is maintained at 86%. This asymmetry reflects the longer opportunity for symptom detection that the pre-flight period provides relative to the flight itself.

**Figure 6:**
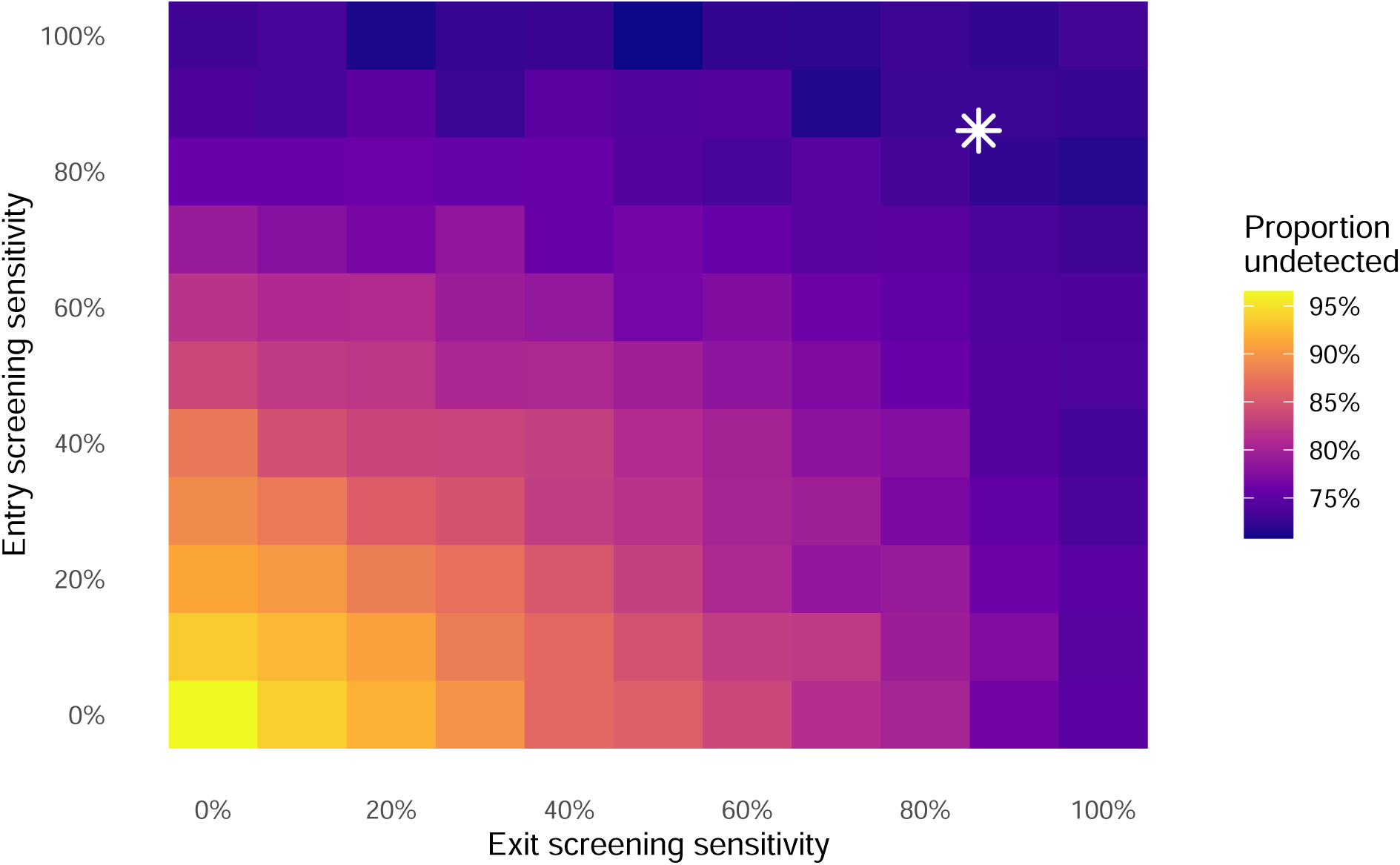
Joint sensitivity analysis: proportion of infected travellers undetected as a function of exit screening sensitivity (x-axis) and entry screening sensitivity (y-axis), for a 12-hour flight with posterior-median BVD natural-history parameters and stable-epidemic exposure distribution. The star marks the primary scenario (86% at both stages). Diagonal dashed lines indicate contours of equal combined coverage.

## 4 Discussion

At 86% sensitivity on a 12-hour international flight, combined syndromic exit and entry screening will fail to detect approximately 73% of infected travellers (95% CrI: 68–77%). The relatively short onset-to-severe-disease interval (mean approximately four days) means that symptomatic travellers progress to detectable illness quickly, yet the incubation period (approximately 7.7 days;^1^) means that most infected travellers will board before symptom onset.

The picture is consistent with prior modelling for Zaire ebolavirus and other viral haemorrhagic fevers, and with the empirical record of Ebola disease outside Africa. Van Zandvoort et al. (2026) reviewed all confirmed Ebola cases presenting outside Africa from 1976 to May 2026 and identified 28 cases in total; of these, only four were latent cases — travellers who passed through exit screening asymptomatically and developed symptoms during or after their journey — giving a crude rate of 0.17 traveller-associated cases per 1,000 reported Ebola cases in Africa since 2000^16^. Notably, these four latent cases occurred among approximately 300,000 travellers screened by CDC-supported exit screening programmes in Guinea, Liberia, and Sierra Leone during the 2014–16 epidemic, and all four were asymptomatic (and hence undetectable) at the point of both exit and entry screening^16^. All four latent cases passed exit screening undetected, consistent with the model prediction that most infected travellers depart during the incubation period. Rapid detection of three of these four cases was ultimately achieved through post-travel self-monitoring protocols rather than airport screening, further underscoring the importance of structured self-monitoring guidance for returning travellers^16^.

In practice, the US Centers for Disease Control and Prevention (CDC) implemented layered exit and entry screening at five designated US airports during the 2014 epidemic, combining temperature checks with structured exposure-risk questionnaires and post-arrival active monitoring for 21 days^17^; our model captures only the syndromic (temperature-based) component of this layered approach, so real-world detection incorporating risk-questionnaire and active-monitoring pathways may exceed our syndromic- screening-only estimates.

Mabey et al. (2014)^10^ estimated that exit screening during the 2014 West Africa ZEBOV epidemic would detect only 7–18% of infected travellers on typical long-haul routes. Assuming 100% exit screening sensitivity, they estimated entrance screening would detect 7% (95% CI: 3–13%) of travellers on a 6.42- hour direct flight and 13% (95% CI: 7–21%) on a 13-hour connecting flight; under the same assumptions, our BDBV model estimates 2.5% (95% CrI: 1.3%–3.9%) and 5.0% (95% CrI: 3.2%–7.1%) respectively. This lower entry detection rate for BDBV, despite its shorter mean incubation period (7.7 vs 9.1 days), reflects its narrower incubation-period distribution (SD 3.5 vs 7.3 days for ZEBOV), leaving fewer rapid-onset cases to transition to symptoms during transit; in both cases, the vast majority of infected travellers (over 93%) still enter undetected regardless of flight duration. Pitman et al. (2005)^18^ similarly showed that entry-only screening for SARS would detect at most one quarter of infected arrivals under 100% sensitivity, falling to fewer than one in ten under realistic instrument sensitivity (30–50%). Gostic et al.^14,15^ demonstrated that screening effectiveness is primarily determined by the ratio of the incubation period to the onset-to-severe-disease interval and the proportion of asymptomatic infection, estimating that even best-case COVID-19 screening would detect a median of only 30% (95% interval 10–53%) of infected travellers during growth. Across the plausible range for BVD, the incubation period remains the dominant source of variation in the undetected fraction, with the asymptomatic fraction secondary given the low rates observed for EVD; only if patients could fly irrespective of symptoms would entrance screening detect a substantial share of cases (43% for ZEBOV, 95% CI 34–53%; 28.3% (95% CrI: 23.5%–33.7%) for BDBV). Taken together, these comparisons across pathogens and outbreaks reinforce that no plausible combination of exit and entry syndromic screening alone is likely to substantially alter the epidemiological trajectory of an EVD-family outbreak; screening is best understood as one layer within a broader response rather than a stand-alone control measure.

This analysis has several limitations. The BVD-specific evidence base for the incubation period is sparse (only three estimates exist^1,12,13^, as described in the Introduction, and the Kratz et al. (2015) estimate rests on only three cases), and the Isiro 2012 line list comprises only 52 cases. As explained in Methods, our propagated credible intervals reflect only within-study sampling uncertainty in the Wamala et al. estimate and likely understate the true uncertainty arising from between-outbreak biological variation or case-ascertainment differences; the incubation-period sensitivity analysis (Figure 4), which spans the wider MacNeil-to-Kratz range, should be consulted alongside the central estimates for this reason.

The 74.3% fever-prevalence estimate from Akilimali et al. (2026)^8^, from which we derive the model’s asymptomatic-equivalent proportion (see Methods), is towards the lower end of pooled estimates for Ebola virus disease more broadly (76%, 95% CI 662̆ 01385%;^9^), so we also report a companion scenario assuming all confirmed cases are febrile (0% asymptomatic-equivalent; see Abstract), which provides a conservative lower bound; the asymptomatic-proportion sensitivity analysis (Figure 4, spanning 0–60%) should be consulted alongside the central estimates for the same reason.

A further set of limitations concerns the natural-history posteriors and travel assumptions. The onset-to-admission Gamma posteriors, whilst appropriately wide, are based on a small sample, and this uncertainty propagates directly into our screening effectiveness estimates. The model also assumes a single uniform flight duration of 12 hours; in reality, travellers from DRC and Uganda take a variety of itineraries with different durations and connections. Our flight-duration sensitivity analysis (Figure 3) shows outcomes vary only modestly across the 10–16-hour range typical of connecting itineraries, so this simplification does not materially affect the conclusions. The assumed 86% screening sensitivity is derived from a thermal-scanning study of influenza border screening^19^, also used in the original COVID-19 screening model^20^; actual EVD detection sensitivity in field conditions may differ substantially given differences in fever thresholds, environmental conditions, and operator training. Because this assumption is not BDBV-specific, the screening sensitivity analysis (Figure 6) should be treated as a primary result, not a secondary check. Post-arrival healthcare-seeking behaviour is also not captured; symptomatic travellers who seek care after arrival represent an additional detection pathway not modelled here.

The model also does not address the specificity of fever-based screening. Fever is a non-specific sign, common to malaria, influenza-like illness, and other endemic febrile conditions in DRC and Uganda; in a population with substantial background febrile-illness prevalence, syndromic screening would generate a considerable number of false positives among uninfected travellers, requiring secondary triage, isolation, or testing capacity that is not modelled here. Nor do we consider the operational or financial cost of running screening programmes (staffing, equipment, and throughput delays); this analysis therefore speaks only to the detection performance of screening, not its overall cost-effectiveness or feasibility at scale.

The model removes travellers from the departing pool once they have progressed to severe disease, treating onset-to-severe-disease as equivalent to the threshold for being too ill to travel. In DRC and Uganda, however, hospitalisation can lag clinical deterioration by hours to days owing to health-system access constraints, geographic barriers, and health-seeking norms. The effective “too ill to travel” threshold is therefore likely to be later than the biological severe-disease onset assumed here, meaning the model may underestimate the proportion of symptomatic travellers who successfully board, an anti-conservative bias in this specific direction.

The model does not account for traveller selection effects. During an active outbreak, some infected individuals may travel specifically to seek care that is unavailable locally, biasing the departing caseload towards symptomatic presentations; conversely, awareness of the outbreak, border communication campaigns, or voluntary self-isolation may deter others, reducing the infected fraction among travellers overall. These pressures operate in opposite directions and are difficult to quantify without data on travel volumes and health-seeking behaviour specific to the outbreak context. The baseline assumption, that infected travellers are drawn at random from the infected population with a uniform exposure-age distribution, is a simplification that may not hold in practice.

Relatedly, the model does not include questionnaire-based exposure risk screening, which forms part of real-world EVD port-of-entry protocols. Since BVD transmission requires direct contact with a symptomatic or deceased case, travellers with known exposure history should, in principle, be detectable by structured risk questionnaires even before fever onset. Incorporating this pathway would increase modelled detection rates above the syndromic-screening-only estimates here.

The simulation model also assumes exposure times are distributed uniformly across the natural history window, approximating a stable (non-growing) epidemic. In an actively growing outbreak, as the current DRC BDBV situation likely is, infection ages among travelling cases skew towards recent exposure, so a greater fraction of departing travellers will be pre-symptomatic and the undetected proportion will be higher than our central estimates suggest. Gostic et al.^14,15^ showed this epidemic-phase effect reduces detection, and our growth-phase sensitivity analysis (Figure 5) demonstrates a similar pattern for BVD: under the initial 90% credible interval for the outbreak’s doubling time (13.8–22.8 days; epiforecasts 2026^2^), the undetected fraction increases relative to the stable-epidemic baseline, and may be higher still under the most pessimistic estimate (doubling time 4.5 days^2^), broadly consistent with the R_0_ of 1.71 estimated by Chamla et al. (2026)^7^. Our central estimates should therefore be interpreted as a lower bound on the undetected fraction during an active growth phase.

Finally, the model conditions on a traveller already being infected and therefore estimates the sensitivity of screening, defined as the probability of detecting an infected traveller, rather than predictive values. Translating these results into positive or negative predictive values, or into expected counts of imported cases, would require assumptions about the prevalence of infection among travellers departing affected areas, which is highly uncertain during an active outbreak and outside the scope of this analysis.

Syndromic airport screening for BVD is unlikely to detect most infected travellers. This conclusion is consistent with the PHEIC temporary recommendations issued by WHO^5^, which advise exit screening in affected countries but explicitly state that entry screening outside the affected region is not considered necessary. Resources directed at airport screening in receiving countries carry an opportunity cost: equivalent investment in contact tracing capacity, isolation infrastructure, and healthcare worker protection in DRC and Uganda would more directly reduce onward transmission at the source. For receiving countries, the more actionable layer of protection is clinician preparedness: healthcare providers in low-incidence settings should consider BVD in the differential diagnosis of febrile illness with a travel history from affected regions, supported by clear referral pathways to high-consequence infectious disease units, pre-agreed diagnostic protocols, and timely communication between national public health authorities and point-of-care clinicians. Complementing syndromic screening with structured post-departure self-monitoring guidance and exposure-history questionnaires—advising travellers to monitor themselves for fever and other EVD symptoms for 21 days and contact healthcare services proactively if symptoms develop—would provide a further safety net for the pre-symptomatic cases that no airport screen can capture, consistent with the successful self-monitoring outcomes described above^16^. Communication of this advice at both departure and destination airports, through travel health notices and airline briefings, could increase uptake.

## Data Availability

All data and code to run the analysis is available at github.com/bquilty25/airport_screening_ebola_bvd.

## Supplementary Material

### 4.1 Figure S1: Entry screening sensitivity by flight duration under 100% exit screening

**Figure 7:**
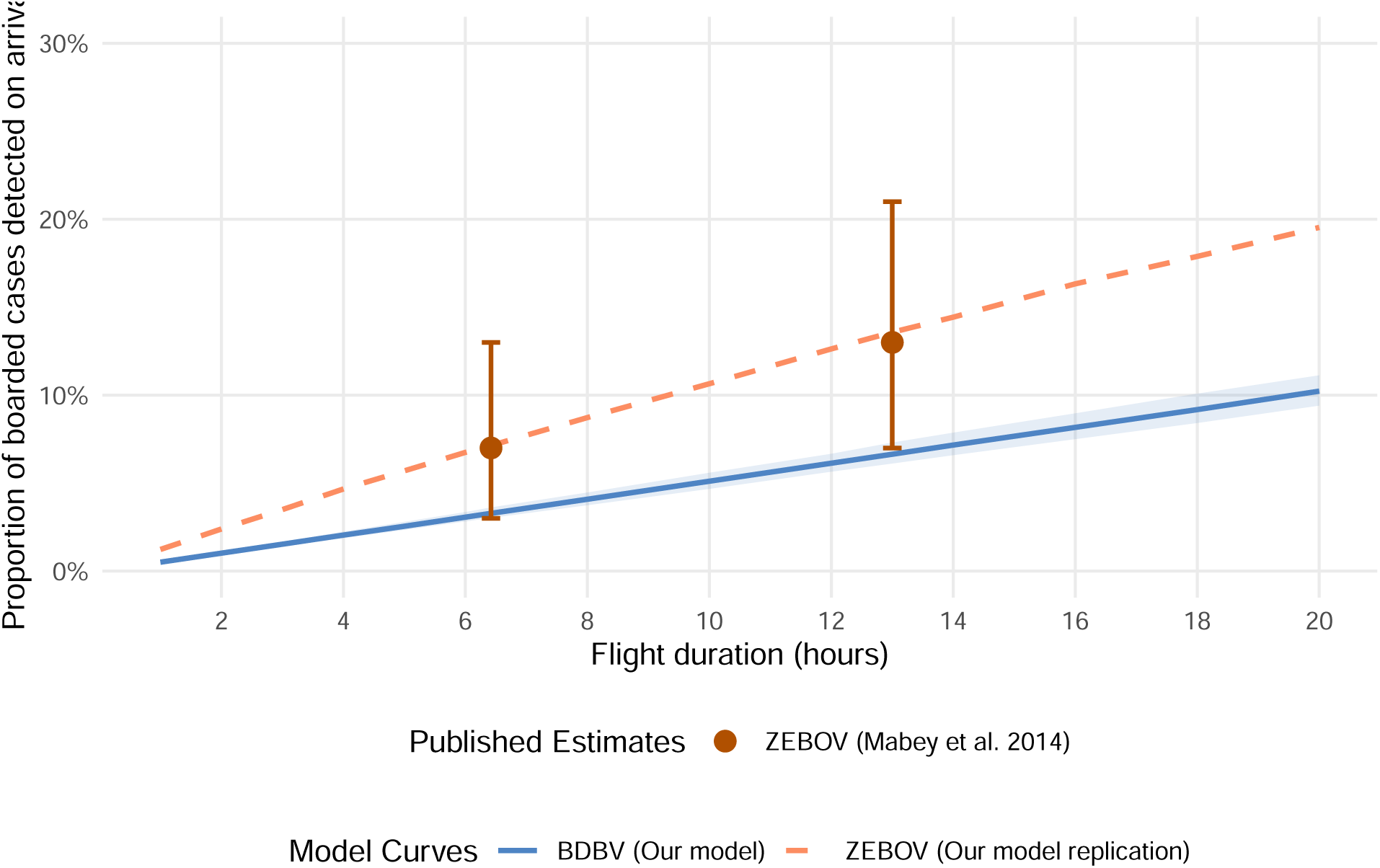
Entrance screening detection probability among successfully boarded travellers as a function of flight duration under a hypothetical 100% sensitive exit screening. The blue ribbon and line show the median and 95% credible intervals for BDBV (propagating Natural History uncertainty and assuming 25.7% asymptomatic-equivalent fraction). The orange line shows the point estimate for ZEBOV (assuming 0% asymptomatic fraction, mean incubation of 9.1 days, and SD of 7.3 days, following Mabey et al. 2014^10^). The point markers with error bars represent the exact direct flight (6.42 hours) and connecting flight (13.0 hours) estimates and 95% confidence intervals from Mabey et al. (2014)^10^ for ZEBOV.

## Acknowledgements

This work was conducted in response to a request for modelling from the UK Foreign, Commonwealth & Development Office. The authors thank Timothy W Russell, Simon R Procter, Joshua Lambert and Nicholas Davies for internal review of the manuscript as part of the CMMID Bundibugyo Working Group. The interactive screening Shiny application and airportscreening R package on which this analysis is based were originally developed for the COVID-19 pandemic response; the application was co-developed with Sam Clifford and the package with Pratik Gupte.

